# “When a thorn comes into your body, you must remove it.” A Socio-Ecological analysis of the Acceptability of Intravaginal Artesunate as Cervical Precancer Treatment in a Phase 1 Clinical trial

**DOI:** 10.1101/2025.10.24.25338731

**Authors:** Annum Sadana, Anagha Guliam, Elizabeth Opiyo, Felix Adundo, Katherine Sorgi, Erum Agha, Chemtai Mungo

**Author notes:** Correspondence: Annum Sadana. **Clinical Trials Registration:** Clinicaltrials.gov NCT06165614.

## Abstract

**Introduction:** Innovative strategies are crucial to meet the World Health Organization’s 90/70/90 cervical cancer elimination targets, and to prevent millions of deaths by 2030. Women in low-and-middle income countries (LMICs) bear the greatest burden of cervical cancer, yet have limited access to cervical precancer treatment. Scalable solutions like self-administered intravaginal therapies can help close this treatment gap. In an ongoing Phase 1 trial of intravaginal artesunate pessaries as primary treatment for cervical intraepithelial neoplasia grade 2 or 3 (CIN 2/3), we evaluated the acceptability of this intervention among trial participants.

**Methods:** 17 participants completed in-depth interviews in their preferred language, focusing on their experiences with artesunate self-administration, including their interactions with the health system, partners, and community while on treatment. In the qualitative study, acceptability was defined as “satisfaction with the product and willingness to use it in the future” as well as “correct and consistent use during the study.” We conducted a thematic analysis using the socioecological framework.

**Results:** Participants in this Phase 1 trial were primarily low-income women, with limited formal education, and the majority were also living with HIV and had been on antiretrovirals for over a decade. Despite minimal prior experience with vaginal self-administration of medication, participants found the pessary acceptable and were highly motivated to get treatment for cervical precancer. Social support, educational interventions, and faith were crucial in enabling participants to complete the treatment protocol successfully.

**Conclusion:** Kenyan women found self-administered intravaginal artesunate for treating CIN2/3 to be highly acceptable. Randomized studies of intravaginal artesunate and other topical therapies in LMICs are needed to evaluate their use in closing the treatment gaps in these settings.

## Introduction

As a preventable disease, cervical cancer is needlessly claiming the lives of women globally. Recognizing this, the World Health Organization (WHO) launched the cervical cancer elimination initiative in 2018^1^. The Global Strategy for Cervical Cancer Elimination defines elimination as reaching a target threshold of 4 cases per 100,000 women-years and established the 90-70-90 targets: 90% of girls fully vaccinated against HPV by age 15, 70% of all women screened for cervical precancer, and 90% of those with cervical disease receiving treatment ^2^. Yet, in the years since the WHO call to action, cervical cancer incidence and death have continued to increase. In 2022, there were 660,000 new cases and 350,000 deaths from the disease globally^3^. Nearly 94% of the deaths in 2022 occurred in low- and middle-income countries, highlighting the inequitable burden of cervical cancer globally. In Eastern Africa, cervical cancer incidence is 40.4 per 100,000 woman-years — ten times the elimination target^3^. Without scale-up of vaccination, screening, and treatment, projections estimate up to 44.4 million new cases of cervical cancer by 2070^4^.

Increases in cervical cancer screening coverage in low and middle-income countries (LMICs) ^5^, will only be effective in preventing cases of cervical cancer if screening is accompanied by high rates of appropriate management of cervical precancer. Current treatment options for cervical precancerous lesions, including cervical intraepithelial neoplasia grades 2 and 3 (CIN2/3), include excisional procedures such as Loop Electrosurgical Excision Procedure (LEEP) and ablative methods like thermal ablation and cryotherapy^6,7^. Despite the introduction of thermal ablation, which can be administered by nurses and does not require anesthesia, access to cervical precancer treatment remains significantly limited due to weak or incomplete completion of treatment cascades, including due to logistical and lab capacity issues for high-quality screening tests, delayed result notification, and a lack of trained health professionals to perform treatment procedures ^8–11^. Rates of appropriate treatment following a positive screening test fall far short of the 90% WHO target, with estimates ranging from 25% in a South African study^9^, 31.8% and 43.3% in a study in Malawi among women with precancer who required excision or cryotherapy, respectively^12^, and from 31.5%^11^ to 50% in two prospective studies in western Kenya^8^.

To achieve the WHO target of 90% of women with cervical precancer receiving treatment globally by 2030, there is an urgent need for scalable, innovative, yet resource-appropriate strategies to close the precancer treatment gap in LMICs. Patient-administered topical therapies pose a promising solution to the issues of access to care faced by women in LMICs. Researchers are investigating two potentially effective therapies for cervical precancer: 5-Fluorouracil (5FU) and artesunate, which are WHO-essential medications. An ongoing randomized trial is evaluating the feasibility of topical, self-administered intravaginal 5FU as an adjuvant to LEEP to reduce CIN2/3 recurrence among WLWH in South Africa (NCT05413811)^13^. In 2020, Trimble et al. published data from a proof-of-concept study of artesunate vaginal inserts (pessaries) for primary treatment of CIN2/3 among HIV-negative women in the United States^14^. In this phase I, dose-escalation study involving 28 women with biopsy-confirmed CIN2/3, the self-administration of three five-day cycles of intravaginal 200 mg artesunate pessaries showed clinically meaningful regression (67%) compared to an expected spontaneous regression rate of 30%^15^. A phase II randomized placebo-controlled trial of artesunate pessaries for the treatment of CIN2/3 among HIV sero-negative women is ongoing (NCT04098744). If randomized efficacy trials support their safety and efficacy, low-cost and accessible self-administered topical agents such as artesunate could be repurposed as primary or adjuvant therapies, increasing treatment access or improving the effectiveness of current cervical precancer treatments and hence strengthening secondary prevention of cervical cancer, especially for women living with HIV (WLWH) in LMICs.

However, it is critical that any new intervention is acceptable and adopted by target populations. The acceptability of intravaginal treatments has previously been studied in the context of topical microbicide candidates for HIV and STI prevention, as well as intravaginal rings for HIV prevention^16^. Initial microbicide acceptability studies focused on product attributes, such as consistency, amount, and applicator use ^17–19^. These studies raised key concerns regarding education on applicator use, partner support, highlighting the need to explore the role of men as “culturally defined decision makers”, and economic determinants of the acceptability of intravaginal treatments^17^. A review of initial studies of microbicides found that these studies lacked an understanding of social processes that would shape microbicide use, including the role of sexual partners, health care providers, and cultural leaders played in acceptability among both men and women^20^. Greene et al (2010) used the socioecological model to examine “inter-personal and contextual factors that shape adherence” to intravaginal microbicides, focusing on interactions with partners and a subset of women working in commercial sex work^21^.

We apply learnings from prior studies of intravaginal treatments for HIV prevention to our analysis of the acceptability of intravaginal treatment for HPV related diseases, including cervical precancer or cervical intraepithelial neoplasia (CIN), which are inextricably linked with the HIV-disease burden in LMICs. Before initiating Phase 1 trials of intravaginal treatments, we surveyed women in western Kenya to understand their perceptions of intravaginal treatments for cervical precancer. We found that of women screened for cervical cancer, 98% would be willing to use a self-administered precancer treatment if available, and 91% believed their partner would support their use of such therapies^22^. We also conducted in-depth interviews with both men and women to understand their perceptions of intravaginal treatments for cervical precancer. Men expressed their willingness to abstain from sex or use condoms if their partner needed such treatment, but were concerned women may be hesitant to ask men to use condoms due to “fear of violence”^23^. In the same nested study, women who participated in focus group discussions (FGDs) were highly receptive to self-administered topical treatments and provided meaningful insights into their potential concerns, ability to negotiate abstinence, and treatment preferences^24^. Furthermore, in a Phase 1 safety trial of intravaginal 5-FU cream conducted in Western Kenya^25^, women found 5-FU acceptable, appreciated the drug’s ease of use, its discreet nature, and the comfort of home application^26^. To our knowledge, there are no qualitative studies that investigate the acceptability of intravaginal artesunate pessaries and describe women’s experiences with these inserts, including contextual factors that impact their adherence.

We aim to fill current gaps in the literature by analyzing in-depth interviews from a Phase I clinical trial of intravaginal artesunate pessaries for CIN2/3, focusing on product, individual, and sociocultural factors that influence acceptability in a clinical trial setting.

## Methods

### Clinical Trial Recruitment and Setting

A Phase 1 trial evaluating intravaginal artesunate for the treatment of CIN2/3 among HIV-seropositive and HIV-seronegative women enrolled 17 participants in Kisumu County, Kenya, and has now completed follow-up (ClinicalTrials.gov NCT06165614). Participants were recruited from western Kenya, a region with a high HIV prevalence (18-22%) compared to the 2024 national average of 3.7% ^27,28^. Seventeen women, aged 18 years or older, with a biopsy-confirmed diagnosis of CIN2/3 were enrolled. Participants had to be non-pregnant, use at least one method of contraception if of childbearing age, and have a CD4 count > 200 cells/µL^29^. Details of the inclusion and exclusion criteria, the participant recruitment process, study visits, and procedures are provided in the published protocol^30^. Before treatment initiation, participants attended at least two education sessions with a study nurse. During these sessions, they learned how to load the pessary into an applicator and used a pelvic model to practice administering the pessary.

The trial’s primary aim is to investigate the safety of self-administered intravaginal artesunate pessaries as a primary treatment for CIN 2/3. Secondary outcomes are adherence, change in lesion size, histologic regression, and acceptability. Participants with regression on biopsy are followed up to 48 weeks. The study was approved by the ethics review boards at the University of North Carolina at Chapel Hill and Amref ESRC Kenya. All participants provided consent for in-depth interviews upon enrollment in the trial.

### Study Team

AS and AG have backgrounds in the social sciences, and AS is a medical student who worked with the study team in Kenya for 6 months while the study was ongoing. EA is a social worker by training and has extensive experience working with qualitative research that incorporates sociocultural dynamics. AS and AG were supervised and trained by EA in qualitative research methodology. AG and EA were not actively involved in the implementation of the Phase 1 artesunate study, maintaining independence from participants and their responses. CM is an obstetrician-gynecologist with extensive clinical trial and research experience in Kenya who oversaw the trial.

### Study Data Collection

Seventeen participants completed in-depth interviews (IDI) after the 8-week artesunate dosing phase. All IDIs were conducted, translated, and transcribed by EAO, a trained social science Research Assistant (RA) with experience conducting qualitative interviews in a clinical trial setting, who was fluent in Luo and Swahili. The IDIs were conducted using an in-depth interview guide specifically developed for this study. The questions focused on participants’ experiences with intravaginal treatments, comfort with administering the drug at home, challenges with the applicator, male partner support, and their reflections on community sentiments around cervical precancer treatment. The interviews were performed in a private room and lasted approximately 60 minutes. Interviews were audio-recorded, and the recordings were transcribed and translated into English. A second RA cross-checked a subset of the transcriptions to confirm that the transcriptions covered the recorded interviews adequately.

### Data Analysis

Three research team members (AS, AG, and EA) followed Braune and Clarke’s six-step process of thematic analysis and generated initial codes from IDI transcripts, following an inductive approach^31^. We read and coded two transcripts, by consensus, selecting codes that summarized content and thematic areas discussed by study participants. After the codebook was created, a third transcript was coded by consensus to refine the codebook and identify any additional concepts. The remaining IDIs were independently coded by AS and AG using Dedoose 10.0.25. Coding discrepancies were reviewed weekly and resolved by consensus. Any new codes and other findings of interest were discussed at weekly meetings between AS, AG, and EA. Once all transcripts were coded, a complete excerpt book was exported and reviewed by AS and AG to generate themes.

### Conceptual Framework

Themes were selected to create a broad framework for the acceptability of intravaginal artesunate within this clinical trial in western Kenya^32^. Themes were created using the socioecological framework, which highlights “individual and social environmental factors as targets for health promotion interventions”^33^. We categorized themes into product, individual, primary-group, health-system, and cultural factors related to the acceptability of intravaginal artesunate.

## Results

Thematic categories for product, individual, interpersonal, health system, and societal factors are summarized in Figure 1.

### Participant Demographics

A majority of participants had a primary or less education, worked in a trade or as a vendor, and were married or living with their partner (Table 1). Most participants earned less than 500 Kenyan Shillings (KSH) per day, did not have electricity at home, nor access to tap water. A majority of participants were also HIV positive, and of those who were HIV positive, most had been on antiretrovirals (ARVs) for over 10 years.

**Table 1:** Demographic Characteristics of 17 participants in a Phase 1 trial of intravaginal artesunate in Kisumu, Kenya.

| Characteristics | Enrolled Participants (N=17)<br>n (%) |
| --- | --- |
| Age (years) Mean (SD) | 39.9 (6.82) |
| Level of education |  |
| Some Primary | 7 (41.2%) |
| Completed Primary | 4 (23.5%) |
| Some Secondary | 5 (29.4%) |
| Completed Secondary | 1 (5.9%) |
| College or higher | - |

| **Occupation** |  |
| --- | --- |
| Salaried work | 1 (5.9%) |
| Business/Trade/Vendor | 10 (58.8%) |
| Farming | 2 (11.8%) |
| Other | 4 (23.5%) |
| **Marital Status** |  |
| Single/Never Married | 1 (5.9%) |
| Married/Living together | 10 (58.8%) |
| Divorced/Separated | 2 (11.8%) |
| Widowed | 4 (23.5%) |
| **Daily Income** |  |
| <Ksh 499 | 14 (8.3%) |
| Ksh >500 – 999 | 3 (17.6%) |
| **Electricity available at home** |  |
| Yes | 7 (41.2%) |
| No | 10 (58.8%) |
| **Tap water available at home** |  |
| Yes | 5 (29.4%) |
| No | 12 (70.6%) |
| **HIV Status** |  |
| HIV-Negative | 2 (11.8%) |
| HIV-positive | 15 (88.2%) |
| **Time since HIV diagnoses** |  |
| < 10 years | 4 (26.6%) |
| > 10 years | 11 (73.3%) |

### Product-level factors

Participants were asked how various product attributes – such as self-insertion, applicator and tampon use, side effects, storage, privacy, convenience, timing, treatment duration, and dosing breaks–impacted their use of the medication (Table 2).

**Table 2.**
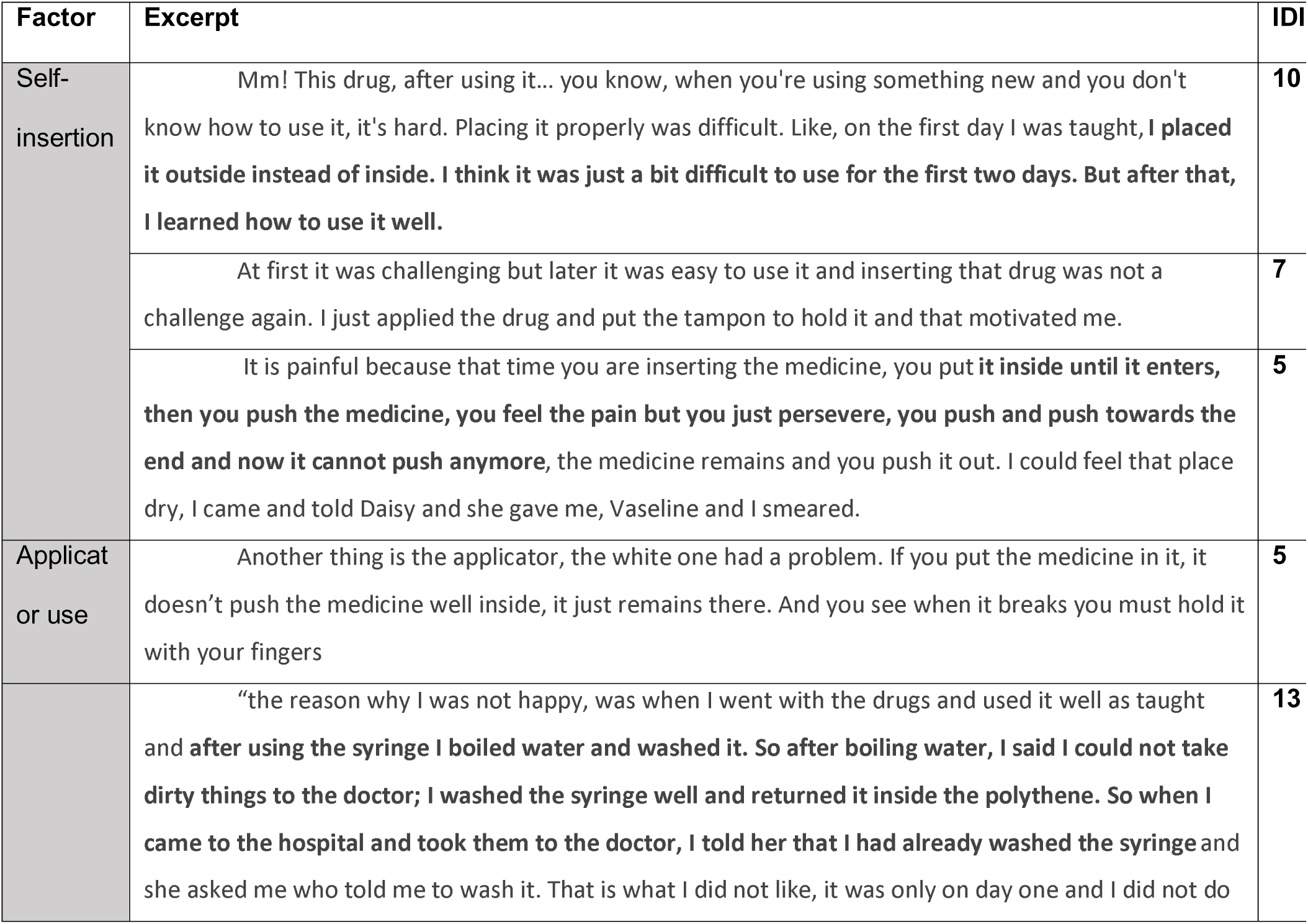
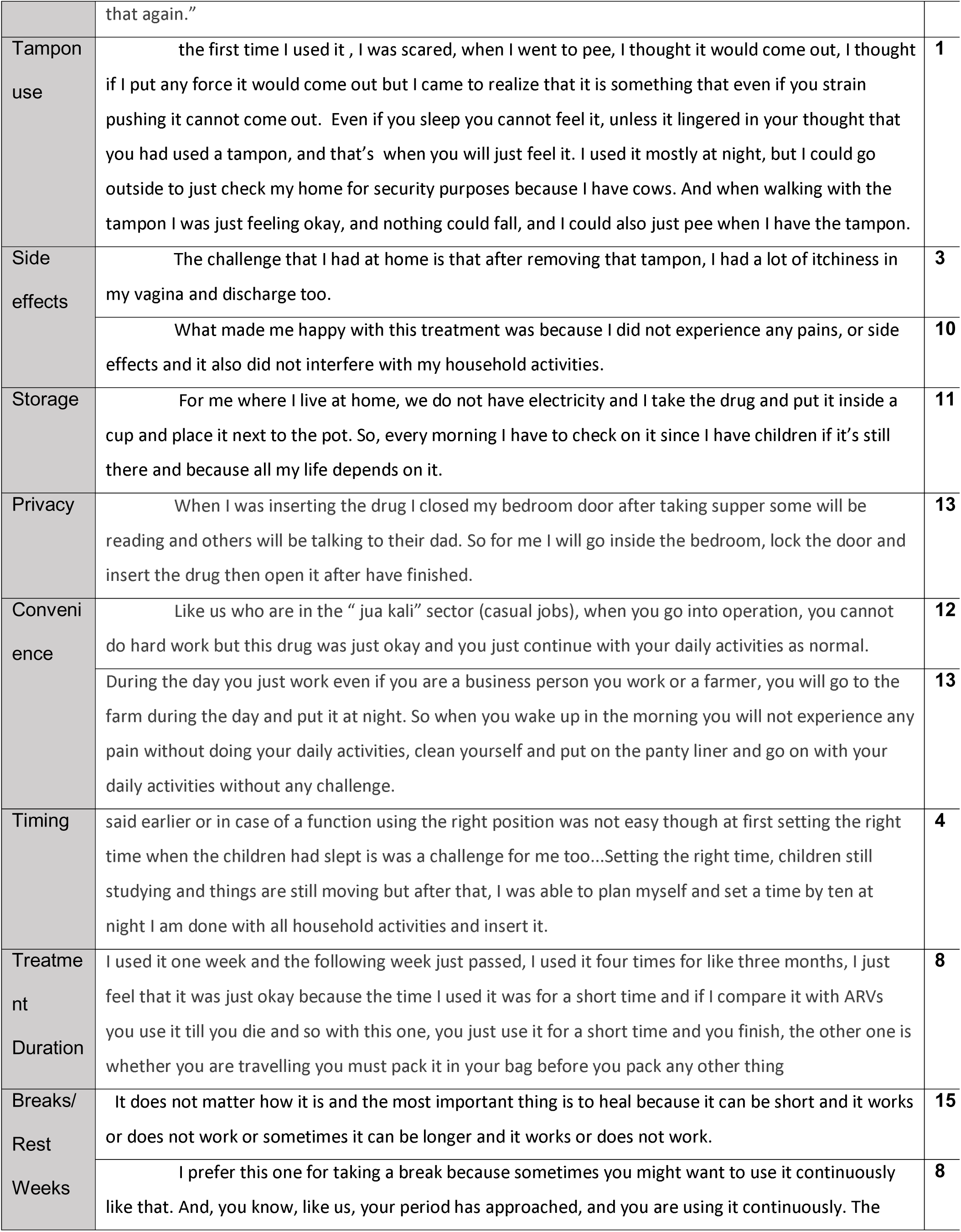
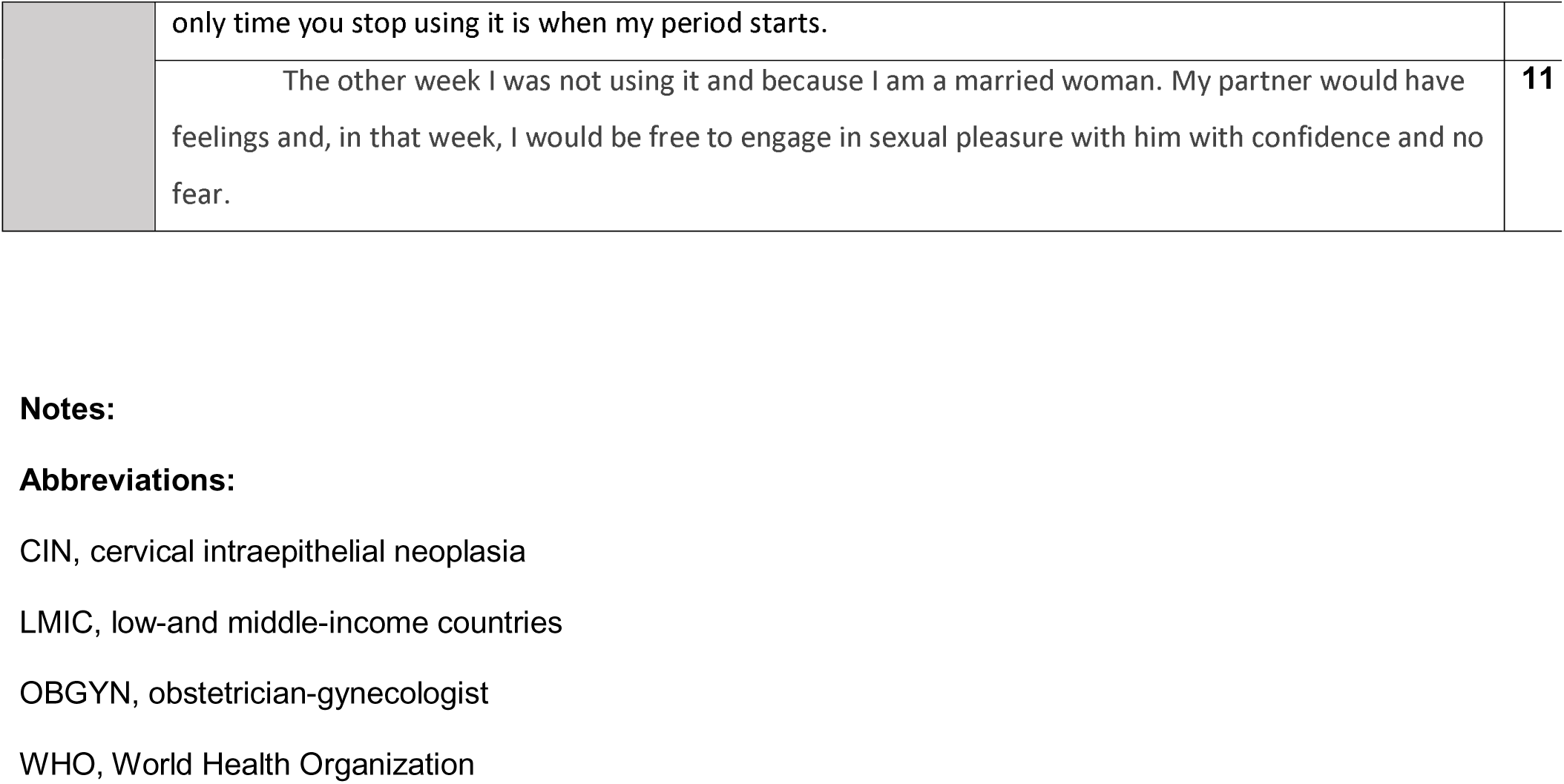
Product level factors related to acceptability of intravaginal artesunate

| Factor | Excerpt | IDI |
| --- | --- | --- |
| Self-insertion | Mm! This drug, after using it... you know, when you're using something new and you don't know how to use it, it's hard. Placing it properly was difficult. Like, on the first day I was taught, I placed it outside instead of inside. I think it was just a bit difficult to use for the first two days. But after that, I learned how to use it well. | 10 |
|  | At first it was challenging but later it was easy to use it and inserting that drug was not a challenge again. I just applied the drug and put the tampon to hold it and that motivated me. | 7 |
|  | It is painful because that time you are inserting the medicine, you put it inside until it enters, then you push the medicine, you feel the pain but you just persevere, you push and push towards the end and now it cannot push anymore, the medicine remains and you push it out. I could feel that place dry, I came and told Daisy and she gave me, Vaseline and I smeared. | 5 |
| Applicat<br>or use | Another thing is the applicator, the white one had a problem. If you put the medicine in it, it doesn't push the medicine well inside, it just remains there. And you see when it breaks you must hold it with your fingers | 5 |
|  | “the reason why I was not happy, was when I went with the drugs and used it well as taught and after using the syringe I boiled water and washed it. So after boiling water, I said I could not take dirty things to the doctor; I washed the syringe well and returned it inside the polythene. So when I came to the hospital and took them to the doctor, I told her that I had already washed the syringe and she asked me who told me to wash it. That is what I did not like, it was only on day one and I did not do | 13 |
|  | that again.” |  |
| Tampon use | the first time I used it , I was scared, when I went to pee, I thought it would come out, I thought if I put any force it would come out but I came to realize that it is something that even if you strain pushing it cannot come out. Even if you sleep you cannot feel it, unless it lingered in your thought that you had used a tampon, and that’s when you will just feel it. I used it mostly at night, but I could go outside to just check my home for security purposes because I have cows. And when walking with the tampon I was just feeling okay, and nothing could fall, and I could also just pee when I have the tampon. | 1 |
| Side effects | The challenge that I had at home is that after removing that tampon, I had a lot of itchiness in my vagina and discharge too. | 3 |
|  | What made me happy with this treatment was because I did not experience any pains, or side effects and it also did not interfere with my household activities. | 10 |
| Storage | For me where I live at home, we do not have electricity and I take the drug and put it inside a cup and place it next to the pot. So, every morning I have to check on it since I have children if it’s still there and because all my life depends on it. | 11 |
| Privacy | When I was inserting the drug I closed my bedroom door after taking supper some will be reading and others will be talking to their dad. So for me I will go inside the bedroom, lock the door and insert the drug then open it after have finished. | 13 |
| Convenience | Like us who are in the “ jua kali” sector (casual jobs), when you go into operation, you cannot do hard work but this drug was just okay and you just continue with your daily activities as normal. | 12 |
|  | During the day you just work even if you are a business person you work or a farmer, you will go to the farm during the day and put it at night. So when you wake up in the morning you will not experience any pain without doing your daily activities, clean yourself and put on the panty liner and go on with your daily activities without any challenge. | 13 |
| Timing | said earlier or in case of a function using the right position was not easy though at first setting the right time when the children had slept is was a challenge for me too...Setting the right time, children still studying and things are still moving but after that, I was able to plan myself and set a time by ten at night I am done with all household activities and insert it. | 4 |
| Treatment Duration | I used it one week and the following week just passed, I used it four times for like three months, I just feel that it was just okay because the time I used it was for a short time and if I compare it with ARVs you use it till you die and so with this one, you just use it for a short time and you finish, the other one is whether you are travelling you must pack it in your bag before you pack any other thing | 8 |
| Breaks/ Rest Weeks | It does not matter how it is and the most important thing is to heal because it can be short and it works or does not work or sometimes it can be longer and it works or does not work. | 15 |
|  | I prefer this one for taking a break because sometimes you might want to use it continuously like that. And, you know, like us, your period has approached, and you are using it continuously. The | 8 |
|  | only time you stop using it is when my period starts. |  |
|  | The other week I was not using it and because I am a married woman. My partner would have feelings and, in that week, I would be free to engage in sexual pleasure with him with confidence and no fear. | **11** |

**Notes:**
**Abbreviations:**
CIN, cervical intraepithelial neoplasia
LMIC, low-and middle-income countries
OBGYN, obstetrician-gynecologist
WHO, World Health Organization

Most participants had not used a self-administered vaginal medication before, and the few participants who had used intravaginal medication primarily used antifungal creams. Most were willing to self-insert and were able to do so correctly and consistently. A few participants initially had challenges with inserting the applicator or tampon into the cervix completely but were able to successfully insert after a few doses (Table 2). One participant asked her partner to help her insert, and another wanted the doctor to insert the pessary for her. A few participants reported insertion-related discomfort or dryness, which resolved after using Vaseline provided by the study, while others noted abdominal pain and cramping from inserting the pessary too deeply. Participants reported challenges with one of the applicators, which did not dispense the full dose of the medication. However, after the applicators were changed, all participants were able to use the applicator for self-insertion successfully. Since none of the participants had previously used a tampon, they initially reported fear or discomfort, at times worrying they would not be able to urinate when using the tampon. However, after several uses and reassurance from the study team, their concerns were alleviated. Participants described side effects they experienced during artesunate use: including headaches, discharge, vaginal itching, abdominal pain, and slight dizziness, but these side effects rarely hindered their ability to do daily activities or carry out their responsibilities.

Overall, most participants found the pessary self-administration process convenient. Most used the pessary at night, although they understood that the pessary could be administered at any time of day if needed. Some participants anticipated challenges with when to administer the pessary, especially when they had multiple obligations, but were able to successfully arrange their schedules to allow them to self-administer the study medication. One participant wanted the study staff to issue reminders nightly at the time of dosing.

Participants had some concerns regarding storage, particularly due to their lack of electricity and the possibility that the pessary might melt at home before use. All participants were able to find a private space to administer the pessary, even if they had families and small children. Some women highlighted that the discreet nature of the medication prevented them from unwillingly unveiling their treatment course and condition to others. The majority of participants did not have strong opinions on the duration of artesunate use and would have been fine with shorter or longer treatment cycles if this was recommended, although a specific maximum length of treatment was not explored. One participant contrasted the short duration of this treatment to her lifelong use of ARVs. Treatment breaks allowed participants to schedule treatment around their menstrual cycles, manage obligations during off-weeks, and maintain sexual activity with their partners. In general, the pessary use was not disruptive to their daily life, and likely contributed to their consistent use.

### Individual factors/participant factors

#### Motivations

Many participants in our study were motivated to either be screened or to join the study after seeing people in their community die from cervical cancer, which contributed to their understanding that treatment for cervical precancer is important. They were highly motivated to heal their cervical precancer.

> “The reason why I was motivated to join the study is how the nurse talked to me, and I knew I was going to be healed.” (13)

Along with their desire to heal, participants understood that others could benefit from the results of the research.

> *“When a thorn comes into your body you must remove it and when I decided to join the study, I was told the treatment was still in research, where they want to know the number of patients that will get help from using the treatment, and I was also encouraged to use it to know the progress of treatment.” (15)*

#### Expectations

Even though this was a Phase 1 trial, and participants understood that they were enrolled in a treatment under research, they wanted artesunate to work for them.

> *“What I can say about that drug is that I had faith it was going to heal me when using it. Like I was also told that in ten people who are using this drug, it can heal seven who will be negative and three positive. So, when I was counselled or educated on how to use this drug and using it, I was counting myself among the seven.” (09)*

The perception that their cervical precancer symptoms were subsiding through the course of the study seemed to reinforce participants’ expectations of the medication’s effectiveness.

> *“So after using the drug, I can say that it is very effective because the abdominal pains ended, discharges ended and I do not have any smell I did not have [side effects] and I used the drug well because before I started using it, I had abdominal and stomach pains and all of them ended…with a lot of faith and trust, I can say that this drug is good even if it’s still on research” (14)*

They chose artesunate because it allowed them to avoid invasive treatment options (such as LEEP) and protected their future childbearing ability by avoiding a possible hysterectomy.

> *“Using this medication will help to heal as you apply the treatment until it ends. So, it’s important if it heals bit by bit and shrinks until it ends because surgery is also so painful.” (15)*

> *“I will have to follow the treatment well and get healed because if I don’t do it, my uterus can be removed, and I will not be able to get more children, meaning that I will not give birth to more children” (11)*

Some participants also perceived the medication as having greater credibility because it had previously been researched in the United States.

> *“they told me there is a study…they have medicine that has been used in western countries for 2 years , and it has helped in western countries. It has been brought to Kenya to see if it can work and so I agreed to try it out and that’s why I came here.” (IDI 2)*

#### Anxieties

In going through the multiple steps of cervical cancer screening, diagnosis of CIN 2/3, and discussion of treatment options, many participants originally thought they had been diagnosed with cancer. Even when they realized it was a precancer diagnosis, rather than cancer, they often felt distress, which manifested in symptoms of stress – both psychological and physiological.

> *“[My partner] found me sleeping on the couch and he asked me “Are you back from the hospital, why are you just sleeping?” I answered am just okay…He went out and came back and still found me still sleeping. …. He asked “why are you sleeping a lot?” I then told him that I have a headache, he then asked. “Why do you have a headache?” I then told him that I went to the hospital and I was screened for cervical cancer, and they diagnosed me with a disease, I have a wound in my cervix.” (01)*

Participants also worried about the progress of the treatment itself. Most participants were concerned about what would happen at the study conclusion, which felt unclear to them. These were their most common questions:

1. How would they know if the precancer lesion size was reduced, and when would they find out the progress of their treatment?

> *“I was eager to know my progress but they told me that the abnormal cells are still in the same position and the last screening examination I will come back for will give me the results to know whether it has reduced or not but that is what is worrying me so much because I feel I have not improved and that is what is in my mind.” (16)*
2. What would happen if the precancer lesion had not gotten smaller?

> *“My question is on my treatment. Like if the samples are taken and they find out that it has spread to another stage. Will I be enrolled again to participate again in the study or what will happen?” (07)*
3. What would happen if the treatment was effective? Would they continue with treatment?

> *“My question is that if this medicine is found to be effective, but I had asked this question and I was told that if I use it and there are no changes, if it spreads, there are other treatments that can be used. So, it is found to be effective and there is positive results, will I continue using these drugs or what will I do,” (06)*
4. Were the doctors being entirely truthful to them about their prognosis?

> *“I would like to treat what I know without hiding what I am going through and also to be told that the stage where I have reached is not easy or to be told how far the progress is and that is what I would like to know.” (15)*

Some participants were also convinced that if they stopped using the medication, the precancer would return.

> *“When I was told to stop, I just knew that things would worsen, because I stopped using the medicine. I wanted to use it until one day that thing disappeared, I like the medicine.” (05)*

> *“[I] asked the doctor who was handling me about what causes cancer. Like the nurse who told me about what causes cancer response was different from the doctor who handled me in the study…I am still not satisfied with the responses that I have received.” (9)*

Even if participants understood the difference between precancer and cancer, the emotional burden was heavy.

#### Skills

To participate in the Phase 1 artesunate trial, some participants had to navigate complex referral systems and advocate for themselves. Participants also actively monitored changes in their body throughout the treatment. Not only did they accurately report side effects to the treatment team, but they also tracked weight gain and mood. Their high level of engagement reflected the value they placed on the treatment; as a few participants noted, it was so important to them, they could not forget it.

> *“We had a paper where we indicated all the challenges we experienced, and we were told to note the time it starts and when it ends, because we were told that the medicine works [similar to] malaria medicine. Like for malaria tablets they work and sometimes you will experience the side effects.” (07)*

### Primary-group factors

#### Partner

Communication with male partners was mixed in our study sample and depended largely on the level of support women expected from their partners. Some women thought it was unnecessary to tell their partners because they didn’t live with them.

> *“When I came to the hospital, he [my partner] was aware that I was sick and when I went back, I told him that I had been told treatment options and the option that I chose was using drugs. The drug I was going to use for five days and then rest for another week. “(11)*

For women with engaged partners, this support was essential to their participation in the study. Partners not only provided emotional encouragement but also offered tangible forms of support, such as reminding participants to insert their medication or reminding them of their clinic appointments, attending clinic appointments with them, and respecting the abstinence periods required by the medication and requested by participants.

> *“Sometimes I could be called and am not reachable and they will call his number. He was free and I also told him that if such a number calls you it’s the [Artesunate] study just receive because that is a reminder for me to go back to the hospital.” (12)*

> *“he asked about the drugs every week and how we will manage it and I told him that I will be given the drugs…. then he said that he was grateful and also wanted me to be healed…I was told to abstain from sex for six weeks and he was okay with it. So, for me, I took three months without sex, and I did not have any challenges with him.” (11)*

Although all participants knew that they could have sex before artesunate treatment, most partnered participants chose to abstain during treatment weeks. Except for one participant’s partner, partners were generally accepting of this abstinence requirement.

“I used to tell him my schedule of using the drug and he did not disturb me on issues related to sexual intimacy but told me to rest until I finished all my drugs.” (13)

Although most partners were generally supportive, some women expressed concern about negative experiences stemming from their partners’ lack of understanding about the need for abstinence.

> *“sometimes you administer the drug and the man does not want to understand, just like I told you that some men are so difficult… like I told my partner that the week I was using this treatment I didn’t have sex but there are some women who went through different challenges.” (12)*

One woman described her partner as ambivalent about her participation in the study, particularly because it was an experimental treatment, but she was able to convince him that she wanted to participate. Overall, partners played a crucial role in women’s participation and completion of the artesunate treatment, with several women noting that they encouraged their partners to attend partner education sessions offered by the study.

> *“For them to be counselled as couples on how to use this treatment because when a woman comes alone, they will have to go back and explain to his partner how the drug is used but some male partners are difficult to deal with and they do not understand which make it difficult to use the drug and sometimes they skip using it… every woman who is joining the study should be accompanied with a male partner and you talk to them, counsel them and they have to sign before their female partners join the study.” (16)*

#### Other forms of support

Participants also received support from their family, including children and siblings. They also received emotional support from other women in support groups in navigating their diagnosis and treatment. One participant even disclosed her treatment to her pastor, who encouraged her to go through with treatment.

> *“in my case I used to store my medication under the pot and all the people living in my house knew the medicine I was using because if they don’t know someone can just throw the medicine thinking that they are not good things that are supposed to be used… I know that even now they are waiting to know the results, the progress, the day I am coming back and they are also happy that their sister can be healed.” (13)*

> *“My third point, I can also say that I shared with a friend about my treatment and she encouraged me that it is good to use the prescribed medication when you are sick to treat that illness and that encouraged me to continue using this treatment. I also did not have any fear when using the drug.” (09)*

### Provider-related factors

#### Patient education

Patient education helped participants understand the difference between precancer and cancer. Particularly, being told that their disease—cervical precancer—is treatable made a sizeable difference in their attitudes toward treatment and gave women hope. Trial participants also received education on biologically female sexual anatomy—namely, the locations of the cervix and vagina—which increased their confidence in self-inserting the study medication and tampons. This instruction proved essential to multiple aspects of protocol compliance, including proper use of vaginal applicators, adherence to menstrual cycle restrictions, and abstinence requirements.

Participants felt they and others *needed* it to complete essential tasks, such as learning how to insert a tampon for the first time.

One participant even shared what she learned with her community:

> *“its not the one taken orally, but it is inserted in the vagina, when they heard that they got so scared, and felt that they would prefer the oral medicine. I later told them that it is a good thing, it is very soft, and there is a specific place where it should rest at, and it goes so smoothly that it doesn’t interfere with anyone. And when it rests at the specific place its supposed to be, even when you push it further, it doesn’t move, then you now pull out the applicator. They were feeling that inserting it with the applicator might be painful, and that is just because they have never used it. It’s not a bad thing it’s a good thing, its never painful." (1)*

#### Interactions with Providers

Participants also greatly appreciated the responsiveness of the study team, particularly their timeliness in addressing concerns such as discomfort with the applicator or any pain or symptom experienced during treatment.

> *“here you don’t waste time, you just come in and you are attended to and then you go back unlike in other places where you come at 8:00 AM and then you leave by 4PM and you reach home by 6PM and what you planned will not work to do other things.” (10)*

Participants also appreciated that the study’s flexibility in accommodating their work schedules and felt well treated well by the clinical team, who provided clear explanations and communicated positively and frequently.

> *“I want to appreciate all the doctors who have attended to me since I started this research. They have handled me well in all my clinic visits…even though my time was very short and after explaining to them my situation they were understanding because I am employed by someone …they allowed me to come here by 3pm or 4pm not to interfere with my source of income and maintain a good relationship with my employer.”(04)*

Participants greatly appreciated receiving updates about their treatment progress and prognosis and felt comfortable with team members who helped them navigate the treatment. However, a few expressed concerns that the team might not always be fully transparent about their treatment progress.

> *“Depending on that opinion, I felt like knowing the progress, and I also had fear in me that maybe there is no progress, but they are afraid to tell me.” (15)*

#### Perceived Credibility

Many participants exhibited an unconditional acceptance of the treatment regimen, largely due to their trust in the trial clinicians’ recommendations.

> *“What I know is that a drug that can help my life, I can use that time I am told to, what matters is that by the end is that I am alive and it has helped me.” (08)*

> *“Yes, because there is no challenge I had when using it, I can say it even gave me a side effect that could hinder me from doing my daily activities, so I can still just use it. If it is working, I can use it.” (08)*

Given the knowledge and resource differences between the study staff and participants, many participants were willing to follow any instructions from the research team in order to receive treatment, including adhering to a longer regimen or different dosing, if recommended, demonstrating motivation to get cured. They were also motivated to meet providers’ expectations and follow instructions.

> *“Because it depends on how the doctor taught me how to use it. Like if they say that we use it continuously, I will just do that” (07)*

### Culture and Environments

#### Cost

A majority of participants lived at or below the World Bank’s international poverty line for lower-middle-income countries (499 KSH); approximately $4 per day^34^, as many worked as farmers or informal laborers. For a majority of the women, the cost of standard cervical precancer treatment, including the cost of transport to a referral hospital, a diagnostic biopsy, and therapeutic LEEP, was challenging, if not impossible to afford. These financial barriers were even greater for women who lived in rural areas or far from the study site.

> *“majority of the women are suffering in the villages and in places like Busia there are so many women who do not go to the hospital. when they are sick they do not go to the hospital, but just buy the drugs which reduces the pain and not healing it. So one day when that disease symptoms will come out, it will come out by force because it has been suppressed by the drugs and harm you. All I can say is that women who are in the villages are suffering.” (13)*

> *“I had gone for screening before, and they diagnosed this problem but I had financial challenges. They prescribed for me a medicine worth four thousand three hundred Kenyan shillings, that is four years back. I had serious financial challenges that I could not even buy medicine. And was suppose to be going for follow ups. I did not go to the clinic because,I didn’t have money to buy the medicine, I thought “ why should I go to the clinic, what will I tell the doctor?” They will prescribe other medicine and I wont be able to buy just like those ones I could not buy.” (02)*

Therefore, many women in the study relied on the free treatment provided by the study to be able to access any cervical cancer treatment.

> *“So I told myself that its God, if I can meet a research study where I do not pay for drugs and transport, that is like I am coming free of charge and just coming to pick my medication then again get something for tea when you reach home which made me so happy.” (13)*

A few women even relied on women’s support groups for *chama*, or mutual aid funds, to be able to access the biopsy to diagnose cervical precancer.

> *“So the lady had to call our leader in Nairobi who sent that money to my M-Pesa number, then I withdrew the money and then I paid.” (16)*

Some women reported that community members offered them alternative medicine for treatment of their cervical precancer, especially due to its lower costs, but they declined it.

#### Religion

To navigate their challenging diagnosis, women relied on religion for hope. Some women prayed immediately before insertion of the pessaries. Others felt grateful that God brought the study to them so they could be treated, and some found solace that God was following them along this journey and would ultimately determine their healing.

> *“When at the clinic and the doctor told me that I was going to get drugs I was so happy but I did not tell her because in my mind I knew that there are no drugs that can cure cancer…I said to myself that God is great …So when on the bed to insert it, that is when I prayed first before administering the drug and I was so happy.” (14)*

## Discussion

Participants in this Phase 1 trial were primarily low-income women with limited formal education, working in informal sectors and living without electricity or tap water; the majority were also living with HIV and had been on antiretrovirals for over a decade. Despite minimal prior experience with vaginal self-administration of medication, participants were able to use the artesunate pessary correctly and found it convenient and discreet for home use. They were highly motivated to treat cervical precancer due to personal and community experiences with cancer, and many perceived artesunate as a less invasive, fertility-preserving alternative to surgical options. Social support—from partners, family, and the study team—as well as strong educational interventions and faith in the research process were crucial in enabling participants to overcome anxieties, adhere to treatment, and complete the protocol successfully.

Participants’ intersectional vulnerabilities are explored in this paper, as they relate to their education level, income level, employment status, gender, and relationship dynamics. The Socio-Ecological model provides a useful lens through which to analyze the acceptance of this intravaginal treatment, as it emphasizes the influence of factors outside the participants’ control, which ultimately impact individual and group acceptance of a novel therapy for cervical precancer.

### Limitations

At the start of the clinical trial, women who enrolled had limited knowledge about cervical precancer and had faced significant challenges in getting a diagnosis in the first place. Their initial hopelessness was converted to hope only by the trial clinician or doctor’s explanation that their condition is treatable. As the interviewer was part of the research team and the interview was conducted in the clinic, the interviewees may have had a desire to meet the interviewer’s expectations. Therefore, some of their responses may reflect this desire, rather than participants’ genuine opinions, a phenomenon known as social desirability bias^35^. To mitigate this, clinical staff left the clinic area during the in-depth interviews.

### Recommendations

Our findings can inform recommendations for future clinical trials and implementation research on the use of intravaginal therapies for cervical precancer.

1. Instructions on Self-Insertion: In this Phase 1 study, women received detailed instructions on self-insertion using a uterine model. However, providing additional guidance on female pelvic anatomy can help clarify the appropriate depth of insertion, which would help ensure that participants feel more confident with intravaginal administration of such therapies.
2. Cancer-related emotional distress and need for social support: A diagnosis of cervical precancer and its treatment can be highly distressing and anxiety-inducing, particularly in settings where cervical cancer is both common and associated with high mortality. As a result, women in this context would benefit from greater education about their condition, such as being shown colposcopy images of cervical abnormalities (when available) and receiving their HPV test results at study exit. Further, peer-support groups for cervical precancer – or integrating HPV/cervical precancer education into existing HIV support groups would help women process the new medical information they are learning as part of their diagnosis. Given the complexity of cervical precancer treatment and high recurrence rates among WLWH, effective prognostication requires skillful counselling and multiple conversations to build a shared understanding of HPV infection and cervical precancer disease course in this population.
3. Partner support: For women with partners, partner support is a crucial determinant of adherence to self-administered intravaginal therapies, especially in the context of abstinence requirements. This need is heightened in contexts where women may have limited agency in negotiating sexual activity due to sociocultural and gender dynamics. Integrating partner-focused education on HPV and cervical precancer can strengthen adherence, enhance treatment outcomes, and mitigate potential relational conflict associated with this diagnosis and intravaginal therapies.
4. Patient education and health provider interactions: The self-insertion of intravaginal medications requires an understanding of female anatomy. Patient education, both in trials and real-world implementation, is essential for participant comfort and adherence. Empowering women with knowledge of reproductive anatomy may also help dismantle the steep hierarchy between patients and healthcare workers.
5. Cost: As artesunate is a WHO essential medication, ensuring access will require it to be provided free of charge or at low-cost if proven efficacious for cervical precancer. Addressing affordability will be critical to achieving widespread access, particularly in LMICs where poverty rates are high.

## Conclusion

In this qualitative study on the acceptability of intravaginal artesunate in Kenya, guided by the socioecological framework, we identified the product, participant, primary group, and sociocultural factors that impact the acceptance of intravaginal treatments. In LMICs where access to cervical precancer treatment is limited, intravaginal therapies – if proven effective- may bridge critical gaps in secondary prevention of cervical cancer. Successful implementation will require careful attention to potential barriers and facilitators to acceptability within the target population at each stage of development. Future randomized trials should continue to explore partner participation, potential impact on sexual function, and the role of HPV-related stigma in shaping women’s experiences with intravaginal treatments for cervical precancer.

## Author contributions

AS: Data curation, formal analysis, visualization, writing of the original draft, writing – review and editing. AG: data curation, formal analysis, writing – review and editing. EA – analysis and supervision, writing – review and editing. EAO conducted, transcribed, and translated interviews. FA contributed to data curation and analysis. KS assisted with project administration. CM conceived of the trial, qualitative interview questionnaire and contributed to writing – review & editing.

## Data Availability

All data produced in the present study are available upon reasonable request to the authors.

## Acknowledgments

The authors are grateful to the participants of the phase 1 study of intravaginal artesunate for their valuable contributions to our understanding of this medication in the context of their lives. We also thank the research teams at KEMRI for their support at the study site, particularly Lizanne Orao for her review of interview transcripts. Thanks also go to Konyin Adewumi for her assistance with coding IDIs. Artesunate vaginal suppositories (pessaries) were provided as in-kind support by Frantz Viral Therapeutics (Mentor, OH).

## Abbreviations

CIN, cervical intraepithelial neoplasia LMIC, low-and middle-income countries OBGYN, obstetrician-gynecologist WHO, World Health Organization

## Ethics Statement

This clinical trial received ethical approval from the following Institutional Review Boards and regulatory authorities: the University of North Carolina at Chapel Hill (USA; approval #23-0902); Amref Health Africa Ethics and Scientific Review Committee (Kenya; approval #P1457/2023); the Kenya Pharmacy and Poisons Board (approval #ECCT/23/09/05), the National Commission for Science, Technology, and Innovation (NACOSTI) (Kenya; approval #NACOSTI/P/24/33045); and the Department of Health Services Kisumu County (approval #GN133VOL.XV/502). Written informed consent was obtained from all study participants before participation.

## References

1. Cervical Cancer Elimination Initiative. Accessed June 24, 2025. https://www.who.int/initiatives/cervical-cancer-elimination-initiative

2. *Global Strategy to Accelerate the Elimination of Cervical Cancer As a Public Health Problem*. 1st ed. World Health Organization; 2020.

3. Bray F, Laversanne M, Sung H, et al. Global cancer statistics 2022: GLOBOCAN estimates of incidence and mortality worldwide for 36 cancers in 185 countries. CA Cancer J Clin. 2024;74(3):229-263. doi:10.3322/caac.21834

4. Simms KT, Steinberg J, Caruana M, et al. Impact of scaled up human papillomavirus vaccination and cervical screening and the potential for global elimination of cervical cancer in 181 countries, 2020–99: a modelling study. Lancet Oncol. 2019;20(3):394–407. doi:10.1016/S1470-2045(18)30836-2

5. Bruni L, Serrano B, Roura E, et al. Cervical cancer screening programmes and age-specific coverage estimates for 202 countries and territories worldwide: a review and synthetic analysis. Lancet Glob Health. 2022;10(8):e1115–e1127. doi:10.1016/S2214-109X(22)00241-8

6. Castle PE, Murokora D, Perez C, Alvarez M, Quek SC, Campbell C. Treatment of cervical intraepithelial lesions. Int J Gynecol Obstet. 2017;138(S1):20–25. doi:10.1002/ijgo.12191

7. World Health Organization. WHO Guideline for Screening and Treatment of Cervical Pre-Cancer Lesions for Cervical Cancer Prevention: Use of Dual-Stain Cytology to Triage Women after a Positive Test for Human Papillomavirus (HPV). Web Annex C. Evidence-to-Decision Framework for Dual-Stain Cytology to Triage Women after a Positive Test for Human Papillomavirus (HPV). World Health Organization; 2024. doi:10.2471/B09020

8. Mungo C, Ibrahim S, Bukusi EA, Truong HHM, Cohen CR, Huchko M. Scaling up cervical cancer prevention in Western Kenya: Treatment access following a community-based HPV testing approach. Int J Gynecol Obstet. 2021;152(1):60–67. doi:10.1002/ijgo.13171

9. Rohner E, Mulongo M, Pasipamire T, et al. Mapping the cervical cancer screening cascade among women living with HIV in Johannesburg, South Africaa. Int J Gynecol Obstet. 2021;152(1):53–59. doi:10.1002/ijgo.13485

10. Cubie HA, Campbell C. Cervical cancer screening – The challenges of complete pathways of care in low-income countries: Focus on Malawi. doi:10.1177/1745506520914804

11. Khozaim K, Orang’o E, Christoffersen-Deb A, et al. Successes and challenges of establishing a cervical cancer screening and treatment program in western Kenya. Int J Gynecol Obstet. 2014;124(1):12–18. doi:10.1016/j.ijgo.2013.06.035

12. Msyamboza KP, Phiri T, Sichali W, Kwenda W, Kachale F. Cervical cancer screening uptake and challenges in Malawi from 2011 to 2015: retrospective cohort study. BMC Public Health. 2016;16(1):806. doi:10.1186/s12889-016-3530-y

13. UNC Lineberger Comprehensive Cancer Center. Acceptability and Feasibility of Combination Treatment for Cervical Precancer Among South African Women Living With HIV. clinicaltrials.gov; 2024. Accessed March 27, 2025. https://clinicaltrials.gov/study/NCT05413811

14. Trimble CL, Levinson K, Maldonado L, et al. A first-in-human proof-of-concept trial of intravaginal artesunate to treat cervical intraepithelial neoplasia 2/3 (CIN2/3). Gynecol Oncol. 2020;157(1):188–194. doi:10.1016/j.ygyno.2019.12.035

15. Trimble CL, Piantadosi S, Gravitt P, et al. Spontaneous Regression of High-Grade Cervical Dysplasia: Effects of Human Papillomavirus Type and HLA Phenotype. Clin Cancer Res. 2005;11(13):4717–4723. doi:10.1158/1078-0432.CCR-04-2599

16. the Microbicide Trials Network 027 Study Team, Bauermeister JA, Golinkoff JM, et al. A Mixed-Methods Study Examining Adherence to and Acceptability of Intravaginal Rings for HIV Prevention: Behavioral Results of MTN-027. AIDS Behav. 2020;24(2):607–616. doi:10.1007/s10461-019-02457-0

17. Joglekar N, Joshi S, Kakde M, et al. Acceptability of PRO2000 Vaginal Gel among HIV un-infected Women in Pune, India. AIDS Care. 2007;19(6):817-821. doi:10.1080/09540120601133576

18. Jones DL, Weiss SM, Chitalu N, Bwalya V, Villar O. Acceptability of Microbicidal Surrogates Among Zambian Women. Sex Transm Dis. 2008;35(2):147–153. doi:10.1097/olq.0b013e3181574dbf

19. Kilmarx PH, Blanchard K, Chaikummao S, et al. A Randomized, Placebo-Controlled Trial to Assess the Safety and Acceptability of Use of Carraguard Vaginal Gel by Heterosexual Couples in Thailand. Sex Transm Dis. 2008;35(3):226. doi:10.1097/OLQ.0b013e31815d6e0d

20. Mantell JE, Myer L, Carballo-Diéguez A, et al. Microbicide acceptability research: current approaches and future directions. Soc Sci Med. 2005;60(2):319–330. doi:10.1016/j.socscimed.2004.05.011

21. Greene E, Batona G, Hallad J, Johnson S, Neema S, Tolley EE. Acceptability and adherence of a candidate microbicide gel among high-risk women in Africa and India. Cult Health Sex. 2010;12(7):739–754. doi:10.1080/13691051003728599

22. Mungo C, Ellis GK, Rop M, Zou Y, Omoto J, Rahangdale L. Perceived acceptability of self-administered topical therapy for cervical precancer treatment among women undergoing cervical cancer screening in Kenya. Adv Glob Health. 2025;4(1):2329438. doi:10.1525/agh.2025.2329438

23. Mungo C, Adewumi K, Ellis G, et al. Men’s perceptions and perceived acceptability of their female partner’s use of self-administered intravaginal therapies for treatment of cervical precancer in Kenya. Ecancermedicalscience. 2024;18:1719. doi:10.3332/ecancer.2024.1719

24. Mungo C, Kachoria AG, Adoyo E, et al. ‘ARVs is for HIV and cream is for HPV or precancer:’ Women’s perceptions and perceived acceptability of self-administered topical therapies for cervical precancer treatment: a qualitative study from Kenya. ecancermedicalscience. 2025;19. doi:10.3332/ecancer.2025.1903

25. Mungo C, Omoto J, Ogollah C, et al. Safety and adherence to self-administered intravaginal 5-fluorouracil cream following cervical intraepithelial neoplasia (CIN) 2/3 treatment among HIV-positive women in Kenya: A phase 1 clinical trial. Int J Gynecol Obstet. 2025;169(2):781–787. doi:10.1002/ijgo.16093

26. Adewumi K, Kachoria AG, Adoyo E, et al. Women’s experiences and acceptability of self-administered, home delivered, intravaginal 5-Fluorouracil cream for cervical precancer treatment in Kenya. Front Reprod Health. 2025;7:1487264. doi:10.3389/frph.2025.1487264

27. National Syndemic Diseases Control Council. HIV 10 Year Progress Report.; 2024.

28. National Syndemic Diseases Control Council. Kenya HIV Estimates Portal. Published online 2025.

29. Sadana A, Omoto J, Sorgi K, et al. Feasibility of intravaginal artesunate as an adjuvant HPV & cervical precancer treatment among women living with HIV in Kenya: Study protocol for a phase II clinical Trial. medRxiv. Preprint posted online June 3, 2025:2025.06.02.25327779. doi:10.1101/2025.06.02.25327779

30. Mungo C, Sorgi K, Hoch C, Tang J, Rahangdale L, Omoto J. WITHDRAWN: Intravaginal artesunate pessaries for treatment of cervical intraepithelial neoplasia 2/3 among HIV-positive and HIV-negative women in Kenya: Study protocol for a pilot trial. medRxiv. Published online December 18, 2024:2024.06.27.24309586. doi:10.1101/2024.06.27.24309586

31. Braun V, and Clarke V. Using thematic analysis in psychology. Qual Res Psychol. 2006;3(2):77–101. doi:10.1191/1478088706qp063oa

32. Braun V, and Clarke V. Using thematic analysis in psychology. Qual Res Psychol. 2006;3(2):77–101. doi:10.1191/1478088706qp063oa

33. McLeroy KR, Bibeau D, Steckler A, Glanz K. An Ecological Perspective on Health Promotion Programs. Health Educ Q. 1988;15(4):351–377. doi:10.1177/109019818801500401

34. June 2025 global poverty update from the World Bank: 2021 PPPs and new country-data. World Bank Blogs. Accessed July 19, 2025. https://blogs.worldbank.org/en/opendata/june-2025-global-poverty-update-from-the-world-bank--2021-ppps-a

35. Bergen N, Labonté R. “Everything Is Perfect, and We Have No Problems”: Detecting and Limiting Social Desirability Bias in Qualitative Research. Qual Health Res. 2020;30(5):783–792. doi:10.1177/1049732319889354

